# Elevated baseline adiponectin levels predict increased risk of progression to tuberculosis among interferon gamma release assay-positive close contacts

**DOI:** 10.64898/2026.05.14.26352472

**Authors:** Gustavo Amorim, Mariana Araujo-Pereira, Kimberly A. Dill-McFarland, Francys A. Rangel, Simon C. Mendelsohn, Marina C. Figueiredo, Marcelo Cordeiro-Santos, Valeria C. Rolla, Fernanda C. Q. Mello, Peter F. Rebeiro, Thomas J. Scriba, Thomas R. Hawn, Bruno B. Andrade, Timothy R. Sterling, the RePORT-Brazil consortium

## Abstract

**Background:** Without tuberculosis preventive therapy (TPT), approximately 5% of individuals infected with *M. tuberculosis* progress to active tuberculosis (TB) disease. Recent studies have identified body mass index (BMI) < 25 kg/m^2^ as a predictor of TB progression, but additional markers are needed to better identify persons at increased risk.

**Methods:** Close contacts of patients with culture-confirmed pulmonary TB were enrolled in the Regional Prospective Observational Research in Tuberculosis (RePORT)-Brazil cohort from 2015 to 2019 and followed for up to 24 months. Analyses were restricted to interferon-γ release assay (IGRA)-positive contacts who did not receive TPT or received <30 days of isoniazid. Prediction models to identify close contacts at increased TB risk were constructed using two complementary approaches: incremental models used BMI as the base predictor and evaluated whether baseline whole-blood transcriptomic signatures, human genetic polymorphism risk scores derived from low-pass whole-genome sequencing, and BMI-related plasma biomarkers improved model discrimination. Agnostic models did not impose BMI in the model and used penalized regression for predictor selection.

**Results:** Among 285 close contacts, 15 (5%) progressed to TB. The model with BMI as unique predictor had a C-index of 0.66 (95% confidence interval [CI] 0.55; 0.77). Adding Rajan5 or Duffy9 transcriptomic signature scores to BMI improved discrimination compared with BMI alone, with C-indices of 0.78 (95% CI 0.62; 0.99) and 0.75 (95% CI 0.61; 0.89), respectively, but did not further improve discrimination after accounting for adiponectin. Adding adiponectin to BMI increased the C-index to 0.80 (95% CI 0.68; 0.91), while adiponectin alone captured most of the discriminatory performance in agnostic models (C-index, 0.80, 95% CI 0.69; 0.91). Genetic risk scores, leptin, and the adiponectin:leptin ratio did not improve model discrimination compared with the BMI-only model. In exploratory post hoc analyses, higher adiponectin was associated with increased risk of progression to TB, with each two-fold increase associated with a higher hazard of TB (HR 2.91, 95% CI 1.73; 4.91, p < 0.001).

**Conclusions:** Baseline adiponectin strongly predicted progression to TB among close contacts and captured most of the discriminatory information contained in epidemiological and transcriptomic variables. Its consistent selection across modelling approaches supports adiponectin as a promising biomarker for TB risk stratification.

## Introduction

*Mycobacterium tuberculosis* infection remains a significant global public health challenge. An estimated 1.7 billion people worldwide are infected with *M. tuberculosis*, the bacterium that causes tuberculosis (TB) disease [1]. Treatment of *M. tuberculosis* infection is highly effective, reducing the risk of TB by 90–95% [2–4], but global coverage of TB preventive therapy remains low [5]. Without treatment, approximately 5–10% of infected persons progress to active TB [6,7], with higher rates among people living with HIV (PLWH) or diabetes [8]. The large reservoir of *M. tuberculosis* infection highlights the need for simple biomarkers to identify persons at increased risk of developing TB, enabling targeted TB prevention and reducing the global burden of TB.

Many studies have sought to identify predictors of progression to TB. However, findings have varied across populations, statistical approaches, and candidate predictors, underscoring the continued need for accurate biomarkers [9]. Some studies aimed to identify risk factors associated with progression to active TB, including markers along the causal pathway to disease [10–12]. Others focused on building prediction models using baseline markers to identify individuals or groups at increased risk of TB progression [13–15].

Recent work has identified lower body mass index (BMI) as a predictor of progression to active TB [13,16,17]. Arriaga et al. (2026) used penalized regression models incorporating multiple demographic and epidemiologic variables and found that lower BMI was a strong predictor of TB progression in an observational cohort in Brazil [13]. In particular, interferon-γ release assay (IGRA)-positive close TB contacts with BMI < 25 kg/m^2^ who did not receive TB preventive therapy were at increased risk of developing active TB. A similar predictive BMI threshold was independently observed by Nguenha et al. (2025) in an analysis of clinical trial data from PLWH in South Africa, Mozambique, and Ethiopia [16]. A recent systematic review including nearly 25 million people showed an inverse association between BMI and TB progression, with risk increasing below 25 kg/m^2^ [17].

Host blood transcriptomic signatures and genetic susceptibility have also been evaluated as predictors of progression to TB [14,18,19]. Mendelsohn et al. (2024) evaluated 19 previously validated host blood transcriptomic signatures and identified three that predicted progression to TB among close TB contacts in Brazil up to 9 months after enrollment [14,20,21]. Although host genetic factors have long been implicated in susceptibility to TB, the major causal genes remain largely unknown, and no genetic variant-based risk predictors of TB progression among exposed contacts have been established.

Since BMI has consistently been associated with TB progression, metabolic profiles may also influence, or reflect, the mechanisms of progression [22]. Although adiponectin has been evaluated in symptomatic versus asymptomatic TB [23,24], its association with progression from *M. tuberculosis* infection to TB disease remains unclear [25].

Building on previous work from our group [13,14], we sought to identify baseline predictors of progression to active TB among genetically heterogeneous close contacts from Brazil. Our aim was to develop robust prediction models, rather than estimate effect sizes or test hypotheses, using demographic, epidemiological, whole-blood transcriptomic, and host genetic polymorphisms. Identifying such markers could help target TB preventive therapy to close contacts at highest risk of progression.

## Methods

### Study population

The study population included close contacts of culture-confirmed pulmonary TB patients enrolled in the Regional Prospective Observational Research in Tuberculosis (RePORT)-Brazil cohort [13,26]. Participants were enrolled across five sites in Brazil located in three Brazilian states: Rio de Janeiro (two sites in Rio de Janeiro and one in Caxias), Bahia (Salvador), and Amazonas (Manaus). Participants were enrolled between June 2015 and June 2019 and followed for up to 24 months for progression to TB disease.

Close contacts were defined as individuals exposed to a culture-positive pulmonary TB patient for more than four hours in one week in the six months prior to TB diagnosis. Contacts were eligible after documented exposure to an active case and had no suspicion of symptomatic TB at time of enrollment, including weight loss. They were evaluated for *M. tuberculosis* infection at baseline (QuantiFERON test) and six months after enrollment (if negative at baseline). Thereafter, follow-up was performed by telephone every six months up to 24 months after enrollment, with in-person visits arranged for participants who reported TB symptoms (e.g., cough, unintended weight loss, fatigue, chest pain), in which a complete evaluation, including CXR, sputum testing (smear, Xpert, culture) and blood/urine collection was performed.

Following Brazilian guidelines, TB preventive therapy was recommended for close contacts living with HIV, those with a positive IGRA result, or children younger than 5 years. The TB preventive therapy regimen was isoniazid for 6–9 months.

Analyses were restricted to close contacts who were IGRA-positive at baseline and either did not receive TB preventive therapy or received <30 days of isoniazid. Further details of the study population have been reported previously [13].

### Data collection

Clinical, laboratory, and sociodemographic data were collected using standardized case report forms in REDCap [27,28] and have been described previously [13]. Contacts with a negative or indeterminate baseline IGRA result underwent repeat testing at month 6. IGRA sample collection, processing, and interpretation were performed according to the manufacturer’s recommendations for the QuantiFERON®-TB Gold In-Tube assay (Qiagen) until May 2017, and the QuantiFERON® -TB Gold Plus assay thereafter.

PAXgene whole blood RNA sample collection at baseline, RNA extraction, transcriptomic signature panel design, and RT-qPCR measurement have been described previously [14]. We considered baseline/enrollment scores for 19 published TB transcriptomic signatures evaluated in that study.

Full genotyping protocol details were recently published and are summarized here. Briefly, 947 pulmonary tuberculosis (PTB) cases and 1807 close contact controls from RePORT-Brazil underwent 1X low-pass whole genome sequencing (WGS) with imputation of whole blood collected at enrollment. Single nucleotide polymorphisms (SNPs) were called using Illumina DRAGEN Germline v4.0.3 and imputed using Illumina DRAGEN Imputation v4.0.3, which implemented GLIMPSE 13 against the 1000 Genomes Project Phase 3 15 (N = 2,489). SNPs were filtered to imputation score > 0.6, minor allele frequency (MAF) > 5%, Hardy-Weinberg equilibrium P > 1E-6, and 0% missingness. Imputed genotypes were then compared to 30X high-pass WGS completed for a subset of individuals (25 controls, 25 cases). SNPs were filtered to those with > 75% concordance with high-pass WGS. Pass-filter, concordant SNPs were then modeled for progression to TB in a genome-wide association study (GWAS) with covariates to account for genetic ancestry (2 principal components), kinship, age, sex, diabetes, HIV status, and smoking history.

Genetic risk scores were derived from the 17 pass-filter, concordant SNPs significantly associated with progression to active TB (P < 5E-8, RISK17). The additive risk model assigned 1 point for each risk allele present. In contrast, the dominant risk model assigned 1 point if at least one risk allele was present, thus treating heterozygous and homozygous carriers equally. Genetic risk scores were then calculated as the sum of points per individual. Additionally, significant SNPs were further verified by probe-based targeted sequencing of a subset of individuals (73 controls, 22 cases), which resulted in ten significant SNPs at 75% concordance across all three methods. These ten triple-concordant SNPs were used to derive additional risk scores in the additive and dominant risk models (RISK10), thus resulting in four risk scores for analysis here.

### Outcome

The primary outcome was progression to active TB among close contacts within 24 months of enrolment. TB progression was classified as microbiologically confirmed, based on a positive Xpert MTB/RIF, Xpert MTB/RIF Ultra, solid culture (Lowenstein-Jensen), or liquid culture (BD MGIT), or clinically diagnosed, based on compatible symptoms, abnormal chest X-ray suggestive of TB, and/or suggestive histology (e.g., caseating or necrotizing granulomas).

### Statistical Analysis

Our study had two aims: first, to evaluate whether adding whole-blood transcriptomic signatures and human genetic risk scores improved prediction beyond BMI alone; and second, to assess whether plasma cytokines and chemokines related to BMI further improved predictive performance. Candidate variables are listed in the **Supplementary Material** and included the original clinical and demographic variables used by [13], with newly available data on 19 TB transcriptomic signatures, four genetic risk scores, adiponectin, leptin, and adiponectin:leptin ratio.

Participant characteristics were summarized as median and interquartile ranges (IQR) for continuous variables, and as counts and frequencies for categorical variables. Continuous variables were compared using the non-parametric Mann-Whitney *U* test for two-group comparisons and the Kruskal-Wallis test for comparisons across more than two groups. Categorical variables were compared using Pearson’s chi-square test. We assumed non-informative censoring. Only baseline data, collected at the of investigation of close contacts, were included in the analysis.

We used two complementary approaches to build and evaluate predictions models. First, we used an incremental approach, building on the BMI-only model from [13] by adding candidate predictors and assessing gains in model discrimination. Second, we used an agnostic approach, fitting penalized regression models to identify the most predictive variables from the full candidate set without forcing BMI into the model. The incremental approach served as the primary analysis, while the agnostic approach assessed the robustness of predictor selection.

#### Incremental prediction models

We started with the BMI-only model from Arriaga et al. (2026) and added candidate predictors one at a time, allowing a maximum of three variables per model to minimize overfitting. We computed Harrell’s concordance index (C-index) for each model to evaluate the added predictive value of candidate biomarkers—genetic risk scores, transcriptomic signatures, adiponectin, leptin, and the adiponectin:leptin ratio—in addition to BMI.

#### Agnostic prediction models

This approach assessed the robustness of predictor selection without prespecifying model structure. We used penalized Cox models with least absolute shrinkage and selection operator (LASSO) regularization to reduce dimensionality and select final predictors [29,30]. Further details are provided in the **Supplementary Material**.

#### Statistical inference

Post hoc analyses measured the association between adiponectin and the hazard of progression to TB. Hazard ratios were interpreted as descriptive associations conditional on variable selection and should therefore be interpreted with caution. We fitted Cox proportional hazard models with adiponectin as the primary covariate, adjusting for BMI. In a secondary analysis, we evaluated this association in a matched dataset, in which TB progressors were matched 1:1 to non-progressors by age (±1 year), sex, BMI (±1 kg/m^2^), and smoking status (current vs non-current). Potential non-linear associations between adiponectin and progression to TB were evaluated using restricted cubic splines with three knots, followed by segmented regression to improve interpretability and identify potential cut-off or inflection points.

Data were complete apart from missingness in some transcriptomic signature scores. This reflected the predefined RT-qPCR quality-control criteria: scores could only be calculated when all required primer-probe Ct values were available, and samples or individual scores with failed primer-probe reactions were excluded. Missingness therefore varied across signatures according to the transcripts included in each model and was assumed to be randomly distributed among participants. Analyses were conducted using complete-case data for each model; no imputation was performed for transcriptomic signature variables due to substantial and potentially informative missingness among TB progressors.

Instead of comparing C-indices from nested models using formal statistical tests, an approach discouraged in contemporary statistical literature [31], we used bootstrap resampling to estimate the distribution of the differences in C-index. We report summary measures from the bootstrap samples, focusing on effect estimation, uncertainty, and consistency across modeling approaches.

All statistical analyses were conducted using the statistical software R (version 4.5.2).

### Ethics Statement

The RePORT-Brazil protocol, informed consent, and study documents were approved by the institutional review boards at each study site (CAAE: 25102412.3.1001.5262) and at Vanderbilt University Medical Center. Participation in RePORT-Brazil was voluntary, and written informed consent was obtained from all participants or their legally responsible guardians. All data were de-identified.

## Results

The study cohort included 285 close contacts who were IGRA-positive at baseline and either did not start TB preventive therapy or received isoniazid for ≤ 30 days. Most participants were female (67%), and the median age was 39 years (IQR 24; 51). Of the 285 contacts, 15 progressed to active TB. As previously reported (see [13]), baseline BMI was lower among TB progressors compared to non-progressors. Demographic, epidemiological, transcriptomic signature, and genetic markers are presented in **Table 1. Table 2** shows the discrimination accuracy of our reference model with BMI as unique predictor (C-index 0.66, 95% confidence interval [CI] 0.55; 0.77).

**Table 1.**
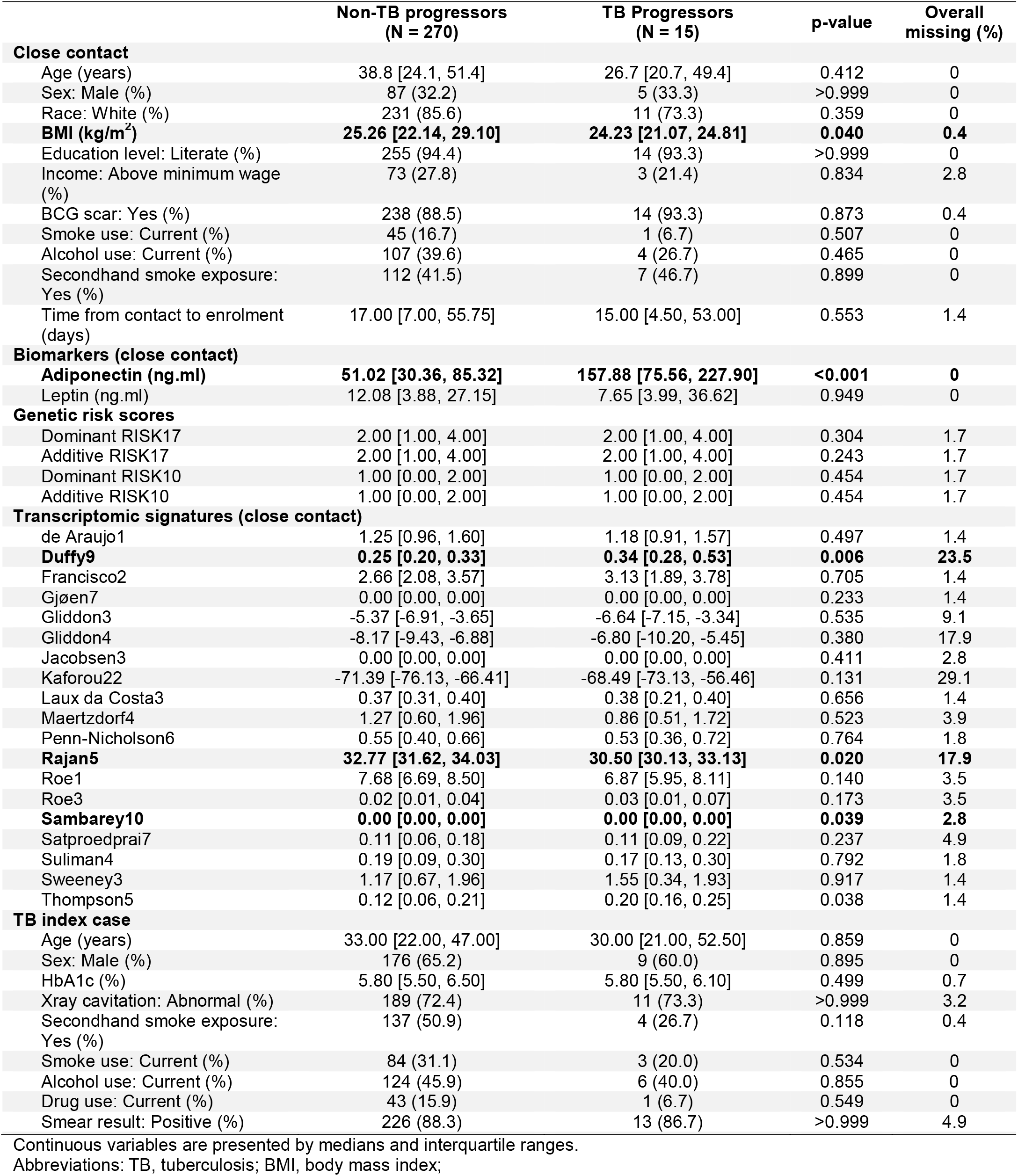
Characteristics of close contacts and TB index cases included in the study.

**Table 2.**
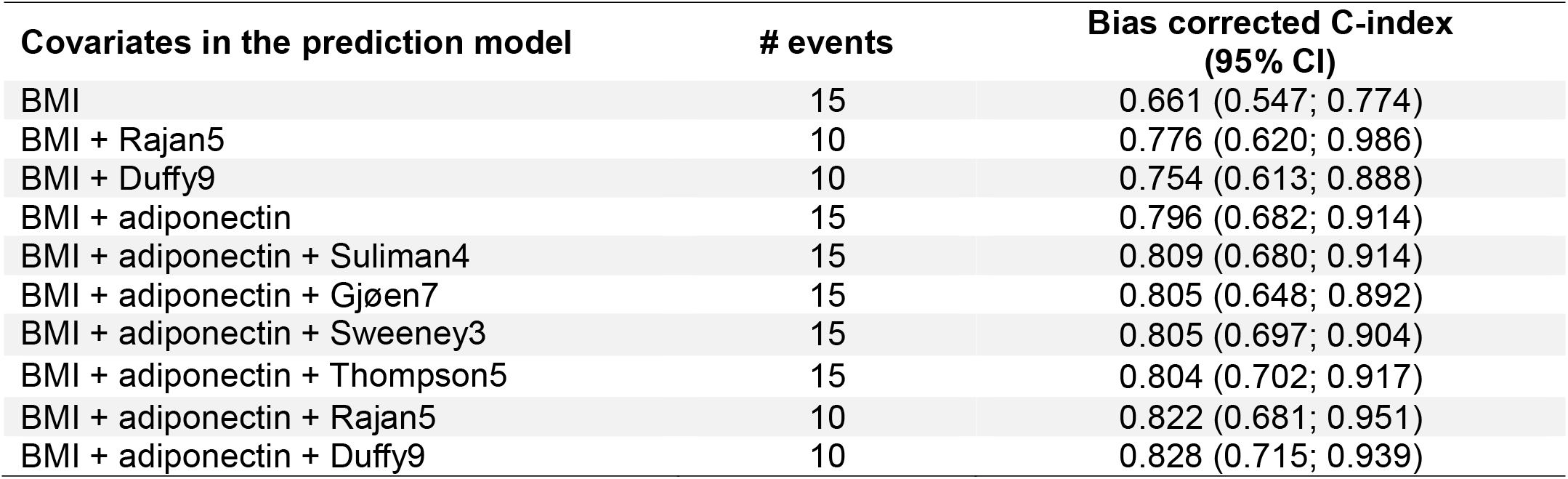
Bias-corrected C-indices for the simplest model with BMI only and other incremental models.

| **Covariates in the prediction model** | **# events** | **Bias corrected C-index**<br>**(95% CI)** |
| --- | --- | --- |
| BMI | 15 | 0.661 (0.547; 0.774) |
| BMI + Rajan5 | 10 | 0.776 (0.620; 0.986) |
| BMI + Duffy9 | 10 | 0.754 (0.613; 0.888) |
| BMI + adiponectin | 15 | 0.796 (0.682; 0.914) |
| BMI + adiponectin + Suliman4 | 15 | 0.809 (0.680; 0.914) |
| BMI + adiponectin + Gjøen7 | 15 | 0.805 (0.648; 0.892) |
| BMI + adiponectin + Sweeney3 | 15 | 0.805 (0.697; 0.904) |
| BMI + adiponectin + Thompson5 | 15 | 0.804 (0.702; 0.917) |
| BMI + adiponectin + Rajan5 | 10 | 0.822 (0.681; 0.951) |
| BMI + adiponectin + Duffy9 | 10 | 0.828 (0.715; 0.939) |

### Transcriptomic signature scores

In univariable analyses, four baseline transcriptomic signature scores were statistically different between TB progressors and non-progressors: Duffy9 (p = 0.006), Rajan5 (p = 0.020), Sambarey10 (p = 0.039), and Thompson5 (p = 0.038; **Table 1**). In the incremental analysis, incrementally adding Rajan5 or Duffy9 to the BMI-only model improved discrimination over the BMI-only model, with C-indices of 0.78 (95% confidence interval [CI] 0.62; 0.99) and 0.75 (95% CI 0.61; 0.89), respectively (**Table 2**). All other transcriptomic signatures did not improve model discrimination (**Supplementary Figure S1**).

Results from the agnostic prediction approach, which did not impose BMI as a baseline predictor, were consistent with the incremental approach. BMI was selected in all models. The only transcriptomic signature scores selected were Gjøeng7, Rajan5, Roe3, Satproedprai7, Thompson5, Duffy9, and Kaforou22 (**Supplementary Table S1**). Overall C-indices were similar to those obtained with BMI alone, although the largest gain was observed when Rajan9 was retained in the model, with a C-index of 0.80 (**Supplementary Table S1**).

### Genetic risk scores

Genetic risk scores were not associated with progression to TB in univariable analysis (**Table 1**). Adding genetic risk scores to the BMI-only model did not improve discrimination in the incremental analysis (**Supplementary Figure S1**). Genetic risk scores were also not selected in the agnostic approach, which did not impose BMI in the model.

### Adiponectin and leptin

Adiponectin was strongly associated with TB progression in univariable analysis, with higher median levels among progressors than non-progressors (p < 0.001; **Table 1**). Adiponectin was inversely correlated with BMI (Spearman correlation ρ = -0.27, p < 0.001), but weakly correlated with transcriptomic signature scores, with the strongest correlation observed for Franscisco2 (Spearman ρ = 0.17; **Figure 1A and C)**. In the incremental analysis, adding adiponectin to BMI improved model discrimination. The BMI-plus-adiponectin model had a C-index of 0.80 (95% CI 0.68; 0.91), compared with 0.66 (95% CI 0.55; 0.77) for BMI alone. The optimism-corrected calibration curve was close to 1, indicating adequate calibration in relative risk, but with large uncertainty (index corrected slope = 0.99, 95% bootstrap CI = 0.50; 1.75). Calibration plot at 2-years after enrollment showed small deviation from observed data (**Supplementary Figure S2**), with large uncertainty due to small number of events. Leptin and the adiponectin:leptin ratio did not improve discrimination (**Table 2**).

**Figure 1.**
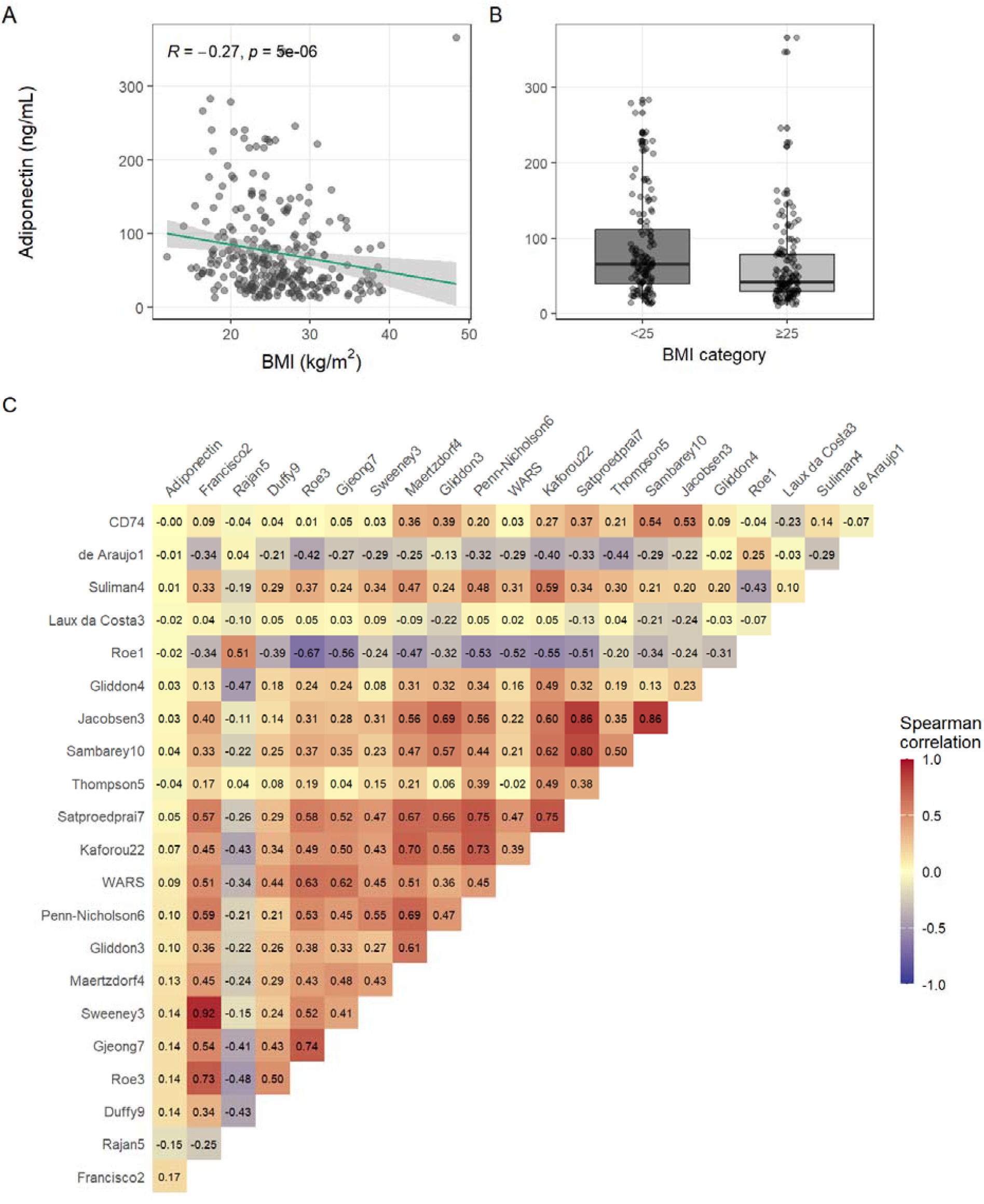
Exploratory analysis showing A) Spearman correlation between adiponectin and BMI, B) boxplot of adiponectin by BMI categories (below or above 25), and C) Spearman correlation matrix of adiponectin and transcriptomic signatures.

Adding adiponectin to BMI increased the C-index by 0.13, with relatively wide uncertainty, likely due to small number of events. To assess the consistency of this improvement, we performed 200 bootstrap resamples and computed the bootstrap distribution of C-index differences. The median bootstrap increase in C-index was 0.15 (IQR 0.11; 0.19) after adiponectin was added (C-index bootstrap difference range: 0.01; 0.39), indicating a consistent increase in predictive performance despite the limited number of events. The agnostic approach further highlighted the importance of adiponectin as a predictor of TB progression. Among all candidate variables, adiponectin was the only predictor of TB progression selected in all ten replications of the penalized Cox models.

### Adiponectin and transcriptomic signatures

We fitted additional incremental models that imposed BMI and adiponectin as the base model, then added each transcriptomic signature individually. After accounting for BMI and adiponectin, transcriptomic signatures did not further improve model discrimination (**Table 2**).

### Risk to TB progression

In exploratory post hoc analyses, log_2_-transformed adiponectin was strongly associated with progression to TB. Each two-fold increase in adiponectin was associated with a 2.92-fold higher hazard of TB progression (HR 2.92, 95% CI 1.73; 4.91, p-value < 0.001). This association remained strong after adjustment for BMI and in sensitivity analysis using a 1:1 matched subset of participants with BMI < 25 kg/m^2^ (**Table 3**).

**Table 3.** Effect of (log2-transformed) adiponectin on progression to active TB in univariable regression, multivariable regression after adjusting for BMI, and on matched analysis restricted to patients with BMI < 25 and < 27.

| **Regression** | **# events** | **Hazard ratio (95% CI)** | **p-value** |
| --- | --- | --- | --- |
| Univariable regression | 15 | 2.92 (1.73; 4.91) | <0.001 |
| Multivariable regression, adjusting for BMI | 15 | 2.82 (1.63; 4.88) | <0.001 |
| Matched analysis, restricted BMI < 25 | 11 | 2.23 (0.96; 5.23) | 0.063 |
| Matched analysis, restricted BMI < 27 | 13 | 2.31 (1.15; 4.67) | 0.019 |

We also explored in post-hoc analysis a potential non-linear relationship between adiponectin and the odds of progression to TB using a segmented regression analysis, similar to the approach used by Arriaga et al. (2026) to identify the BMI inflection point of 25 kg/m^2^. **Supplementary Figure S3** suggested a steeper slope at lower adiponectin levels, with flattening at higher levels. A segmented regression analysis separated by two linear curves was fitted, and a breakpoint was estimated at 221 ng/mL (95% CI 153; 289). Using this data-driven cut-off, 6 of 15 progressors had baseline adiponectin above 221 ng/mL, compared with 9 of 270 non-progressors.

## Discussion

We evaluated the contribution of 19 host blood transcriptomic signatures, genetic susceptibility, and metabolic profiles to identify participants infected with *M. tuberculosis* who are more likely to progress to active TB. We used two complementary modelling approaches using robust penalized regression models with internal validation: an incremental approach building on BMI, and an agnostic approach that did not impose BMI in the prediction model.

Adiponectin consistently emerged as the strongest predictor. Transcriptomic signatures and genetic risk scores added little to no gain in predictive value after accounting for adiponectin. In the agnostic approach, adiponectin was the only predictor selected from the large pool of demographic, epidemiologic, transcriptomic, metabolic, and genetic variables.

Some transcriptomic signatures improved discrimination between progressors and non-progressors to TB in incremental models that adjusted for BMI alone. However, these gains occurred in models with substantial missingness among progressors, raising the possibility of informative missingness, resulting in biased and unreliable estimates. Missingness reduced both the analytic sample size and number of events: for Rajan5, the sample was reduced to 234 close contacts, including 10 progressors, while for Duffy9, the sample was reduced to 218 close contacts, also including 10 progressors. Importantly, these apparent gains in model discrimination, even in these biased samples, were not observed after accounting for adiponectin. This suggests that adiponectin captured much of the predictive signal contained in BMI, transcriptomic signatures, and genetic markers.

Emerging evidence suggests that immunometabolic pathways play a central role in shaping host susceptibility and progression to TB. Adiponectin is involved in metabolic homeostasis and immune regulation, and has been implicated in inflammatory modulation, macrophage function, and systemic metabolic balance. This adipokine may act as a central regulator linking immune activation and metabolic reprogramming during the transition from infection to active disease [32,33].

This study had several limitations. The number of TB progressors was small, limiting the number of predictors that could be included and increasing the risk of overfitting. We sought to minimize overfitting by restricting models to a small number of covariates, but the limited number of events may still have resulted in unstable estimates. Our findings should therefore be interpreted as exploratory and as a comparative evaluation of biomarker classes, rather than as support for immediate model deployment. We were also unable to estimate time since infection, and some contacts may have been exposed to TB earlier in life. In addition, we did not have detailed information on contact type or proximity to the index case. Finally, this was a prediction study focused on risk modelling, and the associations observed should not be interpreted as causal. The underlying biological mechanisms linking adiponectin to risk of TB progression remain unclear and warrant further investigation, including studies of immunological and cellular pathways using complementary single-cell and genetic approaches.

Given the small number of TB progressors, our analyses emphasize parsimony, internal validation, and robustness across modeling strategies. Both modelling approaches showed that BMI was consistently predictive, while adiponectin emerged as the strongest non-BMI predictor and achieved high discrimination between TB progressors and non-progressors as a standalone marker. The convergence between the two approaches supports the interpretation that adiponectin captures a robust signal for predicting progression to active TB.

## Data Availability

All data produced in the present study are available upon reasonable request to the authors

## References

1. Houben RMGJ, Dodd PJ. The Global Burden of Latent Tuberculosis Infection: A Re-estimation Using Mathematical Modelling. PLoS Med 2016; 13:e1002152.

2. Haigh KA, Twabi HH, Boloko L, et al. Efficacy and safety of higher dose rifampicin in adults with presumed drug-susceptible tuberculosis: an updated systematic review and meta-analysis. eClinicalMedicine 2024; 0. Available at: https://www.thelancet.com/journals/eclinm/article/PIIS2589-5370(24)00436-X/fulltext. Accessed 3 January 2025.

3. Sterling TR, Scott NA, Miro JM, et al. Three months of weekly rifapentine and isoniazid for treatment of Mycobacterium tuberculosis infection in HIV-coinfected persons. AIDS 2016; 30:1607–1615.

4. Sterling TR, Villarino ME, Borisov AS, et al. Three months of rifapentine and isoniazid for latent tuberculosis infection. N Engl J Med 2011; 365:2155–2166.

5. Masini E, Mungai B, Wandwalo E. Tuberculosis preventive therapy uptake barriers: what are the low-lying fruits to surmount this? Public Health Action 2020; 10:3.

6. Dheda K, Barry CE, Maartens G. Tuberculosis. Lancet 2016; 387:1211–1226.

7. Targeted tuberculin testing and treatment of latent tuberculosis infection. This official statement of the American Thoracic Society was adopted by the ATS Board of Directors, July 1999. This is a Joint Statement of the American Thoracic Society (ATS) and the Centers for Disease Control and Prevention (CDC). This statement was endorsed by the Council of the Infectious Diseases Society of America. (IDSA), September 1999, and the sections of this statement. Am J Respir Crit Care Med 2000; 161:S221–247.

8. Tornheim JA, Dooley KE. Tuberculosis Associated with HIV Infection. Microbiology Spectrum 2017; 5:10.1128/microbiolspec.tnmi7-0028–2016.

9. Lalvani A, Berrocal-Almanza LC, Halliday A. Predicting progression to active tuberculosis: A rate-limiting step on the path to elimination. PLoS Med 2019; 16:e1002814.

10. Tavitian-Exley I, Kyaw WM, Kang-Yang LL, et al. Risk factors for tuberculosis among close IGRA-negative contacts of persons with infectious tuberculosis in Singapore. International Journal of Infectious Diseases 2024; 147:107166.

11. Asare-Baah M, Lauzardo M, Johnston L, Dominique L, Séraphin MN. Timing and Predictors of Tuberculosis Incidence among Contacts. 2024; :2024.11.02.24316631. Available at: https://www.medrxiv.org/content/10.1101/2024.11.02.24316631v1. Accessed 21 April 2026.

12. Aibana O, Franke MF, Huang C-C, et al. Vitamin E Status Is Inversely Associated with Risk of Incident Tuberculosis Disease among Household Contacts. J Nutr 2018; 148:56–62.

13. Arriaga MB, Amorim G, Figueiredo MC, et al. Body Mass Index and Incident Tuberculosis in Close Tuberculosis Contacts. Clin Infect Dis 2026; 82:e100–e109.

14. Mendelsohn SC, Andrade BB, Mbandi SK, et al. Transcriptomic Signatures of Progression to Tuberculosis Disease Among Close Contacts in Brazil. J Infect Dis 2024; 230:e1355–e1365.

15. Wolde HM, Kebede W, Yewhalaw D, Abebe G, Bae Y, Lee SW. Machine learning approaches to predict the risk of tuberculosis among household contacts of index TB patients in Central Ethiopia. Sci Rep 2026; 16:10457.

16. Nguenha D, Ndebele F, Saavedra B, et al. BMI as a predictor of progression from TB infection to active TB in PLHIV. Int J Tuberc Lung Dis 2025; 29:54–59.

17. Saunders MJ, Cegielski JP, Clark RA, Houben RMGJ, McQuaid CF. Body mass index and tuberculosis risk: an updated systematic literature review and dose–response meta-analysis. Int J Epidemiol 2025; 54:dyaf154.

18. Gupta RK, Turner CT, Venturini C, et al. Concise whole blood transcriptional signatures for incipient tuberculosis: a systematic review and patient-level pooled meta-analysis. Lancet Respir Med 2020; 8:395–406.

19. Mulenga H, Zauchenberger C-Z, Bunyasi EW, et al. Performance of diagnostic and predictive host blood transcriptomic signatures for Tuberculosis disease: A systematic review and meta-analysis. PLoS One 2020; 15:e0237574.

20. Rosain J, Kong X-F, Martinez-Barricarte R, et al. Mendelian susceptibility to mycobacterial disease: 2014-2018 update. Immunol Cell Biol 2019; 97:360–367.

21. Hill AV. The genomics and genetics of human infectious disease susceptibility. Annu Rev Genomics Hum Genet 2001; 2:373–400.

22. Shibata R, Ouchi N, Ohashi K, Murohara T. The role of adipokines in cardiovascular disease. J Cardiol 2017; 70:329–334.

23. Keicho N, Matsushita I, Tanaka T, et al. Circulating levels of adiponectin, leptin, fetuin-A and retinol-binding protein in patients with tuberculosis: markers of metabolism and inflammation. PLoS One 2012; 7:e38703.

24. Santucci N, D’Attilio L, Kovalevski L, et al. A multifaceted analysis of immune-endocrine-metabolic alterations in patients with pulmonary tuberculosis. PLoS One 2011; 6:e26363.

25. Soh AZ, Tan CTY, Mok E, et al. Adipokines and the risk of active TB: a nested case-control study. Int J Tuberc Lung Dis 2021; 25:31–35.

26. Arriaga MB, Amorim G, Queiroz ATL, et al. Novel stepwise approach to assess representativeness of a large multicenter observational cohort of tuberculosis patients: The example of RePORT Brazil. Int J Infect Dis 2021; 103:110–118.

27. Harris PA, Taylor R, Minor BL, et al. The REDCap consortium: Building an international community of software platform partners. Journal of Biomedical Informatics 2019; 95:103208.

28. Harris PA, Taylor R, Thielke R, Payne J, Gonzalez N, Conde JG. Research electronic data capture (REDCap)—A metadata-driven methodology and workflow process for providing translational research informatics support. Journal of Biomedical Informatics 2009; 42:377–381.

29. Tibshirani R. The lasso method for variable selection in the Cox model. Stat Med 1997; 16:385–395.

30. Hossain MdB, Sadatsafavi M, Johnston JC, Wong H, Cook VJ, Karim ME. LASSO-Based Survival Prediction Modeling with Multiply Imputed Data: A Case Study in Tuberculosis Mortality Prediction. The American Statistician :1–12.

31. Harrell, FE. Regression Modeling Strategies: With Applications to Linear Models, Logistic and Ordinal Regression, and Survival Analysis. Cham: Springer International Publishing, 2015. Available at: https://link.springer.com/10.1007/978-3-319-19425-7. Accessed 1 June 2025.

32. Keicho N, Matsushita I, Tanaka T, et al. Circulating Levels of Adiponectin, Leptin, Fetuin-A and Retinol-Binding Protein in Patients with Tuberculosis: Markers of Metabolism and Inflammation. PLOS ONE 2012; 7:e38703.

33. van Crevel R, Karyadi E, Netea MG, et al. Decreased Plasma Leptin Concentrations in Tuberculosis Patients Are Associated with Wasting and Inflammation. J Clin Endocrinol Metab 2002; 87:758–763.

