## Supplementary Material for "Elevated baseline adiponectin levels predict increased risk of progression to tuberculosis among interferon gamma release assay-positive close contacts"

**Supplementary Table S1.** Optimism-corrected C-indices for each of the 19 models, as well as variables selected into the final prediction models after LASSO from a set of candidate predictors from all epidemiological and demographic variables, plus one signature.

| Signature added as candidate variable | Variables selected | Optimism corrected C-index |
|---|---|---|
| Suliman4 | • BMI<br>• Race<br>• Secondhand smoke exposure (index) | 0.712 |
| de Araujo1 | • BMI<br>• Race<br>• Secondhand smoke exposure (index) | 0.747 |
| Francisco2 | • BMI<br>• Race<br>• Secondhand smoke exposure (index) | 0.747 |
| Gjøen7 | • BMI<br>• Gjøen7<br>• Secondhand smoke exposure (index) | 0.722 |
| Gliddon3 | • BMI<br>• Secondhand smoke exposure (index) | 0.694 |
| Jacobsen3 | • BMI<br>• Secondhand smoke exposure (index) | 0.702 |
| Laux da Costa3 | • BMI<br>• Race<br>• Secondhand smoke exposure (index) | 0.707 |
| Maertzdorf4 | • BMI<br>• Race<br>• Secondhand smoke exposure (index) | 0.710 |
| Penn-Nicholson6 | • BMI<br>• Race<br>• Secondhand smoke exposure (index) | 0.714 |
| Rajan5 | • BMI<br>• Race<br>• Rajan5 | 0.801 |
| Roe1 | • BMI | 0.675 |
| Roe3 | • BMI<br>• Roe3 | 0.670 |
| Sambarey10 | • BMI<br>• Sambarey10 | 0.650 |
| Satproedprai7 | • BMI<br>• Satproedprai7<br>• Secondhand smoke exposure (index) | 0.701 |
| Sweeney3 | • BMI<br>• Race<br>• Secondhand smoke exposure (index) | 0.707 |
| Thompson5 | • BMI<br>• Secondhand smoke exposure (index)<br>• Thompson5 | 0.727 |
| Gliddon4 | • BMI<br>• Race | 0.764 |
| Duffy9 | • BMI<br>• Duffy9<br>• Race | 0.787 |
| Kaforou22 | • BMI<br>• Kaforou22 | 0.720 |

| |
|---|
| • Secondhand smoke exposure (index) |

Note: Differences in C-indices across models with same predictors are due to Monte Carlo variability from boostrapping used for internal validation.

**Supplementary Figure S1**. Bias-correct C-index, after 200 bootstrap replications, for all incremental prediction models, building upon BMI including one predictor at a time. The 95% confidence intervals are shown by the dashed lines.

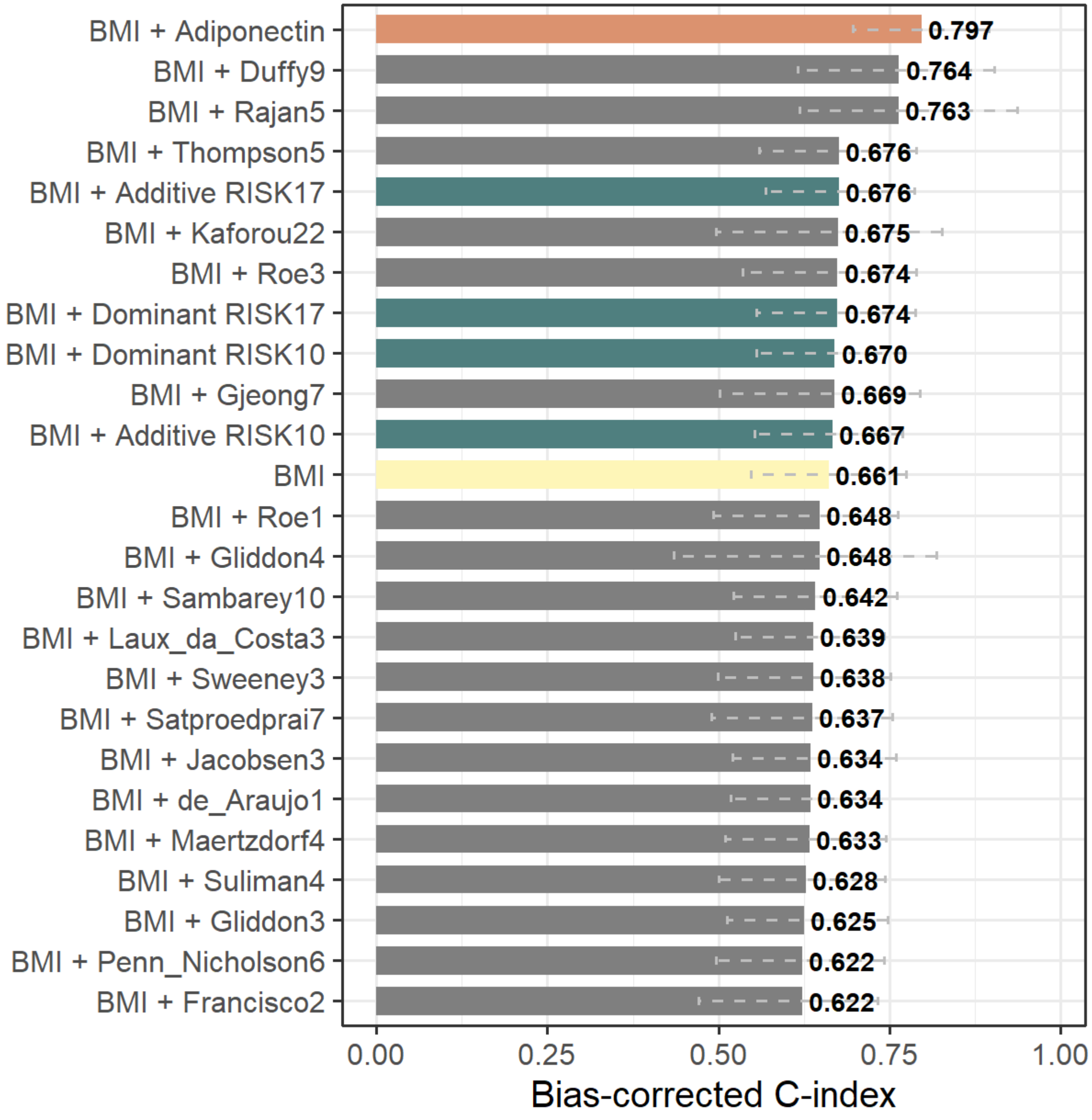

**Supplementary Figure S2.** Calibration plot and optimism-correct calibration slope, evaluated at 720 days after enrollment.

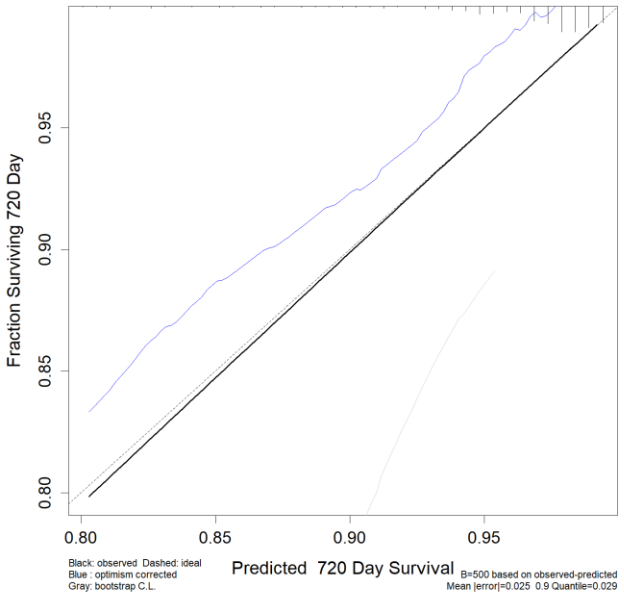

**Supplementary Figure S3.** Panel A shows the non-linear effect of BMI and adiponectin in the odds of progressing to active TB and panel B shows segmented regression model with the optimal data-driven cut-off/breakpoint value.

A)

B)

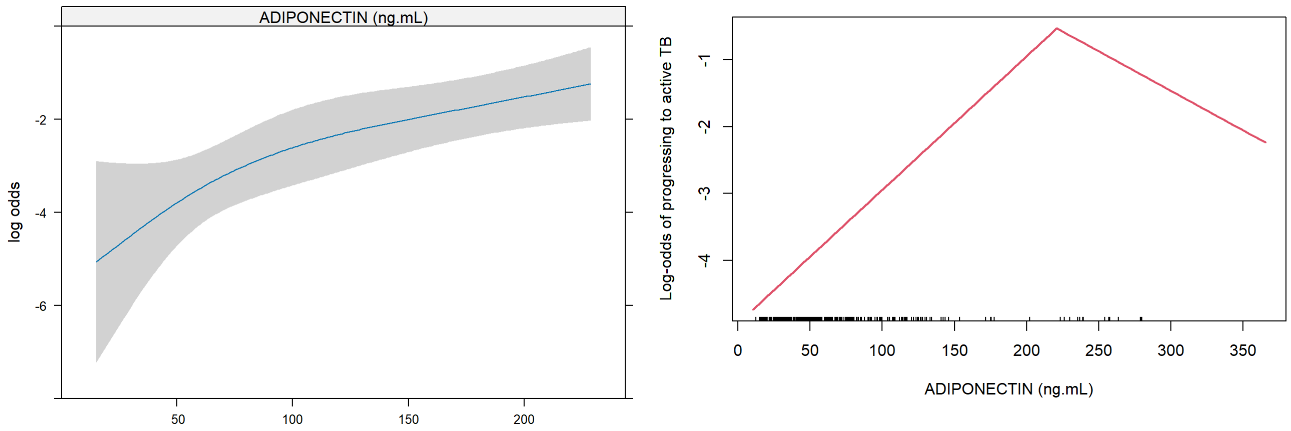

**Agnostic prediction models:**

We also took an agnostic approach that does not impose BMI to be in the final prediction models. These models were constructed following the same multi-step variable selection strategy used in Arriaga et. al. (2026) [1]. First, an empirical review excluded variables with high percentage of missingness (e.g., CAGE questionnaire) or variables with no variability, i.e., variables that were exclusively observed among progressors or controls, to increase model stability. Penalized Cox models were used to reduce dimensionality and select predictors for the final model. We implemented the least absolute shrinkage and selection operator (LASSO).

Because the endpoint was time-to-event, we used the partial log-likelihood as the loss function for both LASSO [2,3]. Penalty/regularization parameters (e.g., λ) were selected via three-fold cross-validation, minimizing the cross-validated partial likelihood deviance. Folds were constructed by randomly splitting the data into three distinct groups of approximately equal size, with outcome-stratification to ensure at least five events per fold. Model discrimination was quantified using Harrell's concordance index (C-index), a survival analogue of the area under the receiver operating characteristic curve (AUC) [4,5]. We focused on optimism correct C-indexes. Due to low number of events, the entire procedure (cross-validation, variable selection, and internal validation) was repeated 10 times and results were averaged across replications.

We decided to evaluate each transcriptomic score separately because of the varying degree missingness: while some were present for all records, other signatures were missing for key participants, including progressors. Using all scores combined would reduce the number of events to eight, nearly a 50% reduction in the initial counts. We constructed 19 penalized regression models using baseline epidemiological, biomarkers, and genetic variables plus one transcriptomic score as candidate variables. For each one of 19 models, we report on the set of variables selected after LASSO and the corresponding Harrell's C-index.

**Supplementary Table S1** shows the final variables for each of the 19 penalized regression models, using the agnostic approach discussed earlier where all epidemiological and demographic variables plus one transcriptomic score for each model are included as candidate variables. Results are displayed for LASSO, using the deviance loss function. Even though BMI was not forced to be in the prediction models, it was consistently selected across all methods and loss functions. For most models, the corrected C-index was around 0.70, the value obtained when only epidemiological variables were considered. However, we noticed a significant increase in C-index after including Rajan5 as a candidate variable, which ended up being selected into the final model, with the C-index increasing from 0.80. It is important to highlight that this result from Rajan5, as well as those for Duffy9, Gliddon4, and Kaforou22 may not be generalizable: the large observed missingness could be informative, leading to a biased sample that is not representative of the study cohort.

### References


1. Arriaga MB, Amorim G, Figueiredo MC, et al. Body Mass Index and Incident Tuberculosis in Close Tuberculosis Contacts. Clin Infect Dis **2026**; 82:e100–e109.

2. Tibshirani R. The lasso method for variable selection in the Cox model. Stat Med **1997**; 16:385–395.

3. Tay K, Simon N, Friedman J, Hastie T, Tibshirani R, Narasimhan B. Regularized Cox Regression. 2025; Available at: https://glmnet.stanford.edu/articles/Coxnet.html. Accessed 5 May 2025.

4. Harrell , FE. Regression Modeling Strategies: With Applications to Linear Models, Logistic and Ordinal Regression, and Survival Analysis. Cham: Springer International Publishing, 2015. Available at: https://link.springer.com/10.1007/978-3-319-19425-7. Accessed 1 June 2025.

5. Harrell FE, Lee KL, Mark DB. Multivariable prognostic models: issues in developing models, evaluating assumptions and adequacy, and measuring and reducing errors. Stat Med **1996**; 15:361–387.
